# Implicit bias in Critical Care Data: Factors affecting sampling frequencies and missingness patterns of clinical and biological variables in ICU Patients

**DOI:** 10.1101/2024.06.09.24308661

**Authors:** Junming (Seraphina) Shi, Alan E. Hubbard, Nicholas Fong, Romain Pirracchio

## Abstract

The presence of missing values in Electronic Health Records (EHRs) is a widespread and inescapable issue. Publicly available data sets mirror the incompleteness found in EHRs. Although the existing literature largely approaches missing data as a random phenomenon, the mechanisms behind these missing values are often not random with respect to important characteristics of the patients. Similarly, the sampling frequency of clinical or biological parameters is likely informative. The possible informative nature of patterns in missing data is often overlooked. For both missingness and sampling frequency, we hypothesize that the underlying mechanism may be at least consistent with implicit bias.

To investigate this important issue, we introduce a novel analytical framework designed to rigorously examine missing data and sampling frequency in EHRs. We utilize the MIMIC-III dataset as a case study, given its frequent use in training machine learning models for healthcare applications. Our approach incorporates Targeted Machine Learning (TML) to study the impact of a series of demographic variables, including protected attributes such as age, sex, race, and ethnicity on the rate of missing data and sampling frequency for key clinical and biological variables in critical care settings. Our results expose underlying differences in the sampling frequency and missing data patterns of vital sign measurements and laboratory tests between different demographic groups. In addition, we find that these measurement patterns can provide significant predictive insights into patient outcomes. Consequently, we urge a reevaluation of the conventional understanding of missing data and sampling frequencies in EHRs. Acknowledging and addressing these biases is essential for advancing equitable and accurate healthcare through machine learning applications.

## 1 Introduction

The digitization of health records through Electronic Health Records (EHRs) has supplanted traditional paper-based systems, thereby centralizing patient-specific information in an electronic medium. Over the past decade, several deidentified EHR datasets were made available to the public, mainly for research purposes. Notable examples include the MIMIC Database (Johnson et al. [2016]), PCORnet(Fleurence et al. [2014]), I2B2 Data (Uzuner et al. [2011]), and the COVID-19 Research Database. The composition of data in publicly accessible datasets is indicative of what is available in EHRs. These empirical data from the real world not only mirror the way care is provided but also inherently incorporate potential biases in how variables are measured. These biases, if not rigorously identified and accounted for, can propagate through analyses.

The advent of large-scale, accessible EHR databases has led to a surge in studies to improve healthcare delivery by identifying patient phenotypic cohorts (Shivade et al. [2014]), developing risk prediction models (Goldstein et al. [2017]), and enriching our understanding of risk factors in relation to health outcomes (Adler and Stead [2015]). However, a significant obstacle in this endeavor is the widespread occurrence of missing values within EHRs. Previous research by Kharrazi et al. [2014], Wells et al. [2013], Botsis et al. [2010], Beaulieu-Jones et al. [2018, 2017] has emphasized the necessity of addressing the issue of missing values and has recognized that incomplete data is often related to clinical practice, rather than being distributed randomly. Missing values may occur for some of the following reasons: data was measured but not recorded, the patient’s condition made measurement impossible, or healthcare professionals had no intention of taking the measurement.

Existing methodologies to mitigate the impact of missing values predominantly include imputation techniques (Sharafoddini et al. [2019]), Inverse Probability Weighting (IPW) (Seaman and White [2013]), ignoring observation with missing data in complete case analysis (CCA) and available case analysis (ACA). Specifically, CCA involves the exclusion of any data rows containing missing values or outliers, thus limiting the analysis to only fully observed cases. ACA, conversely, utilizes all available data specific to each analytic question and is also known as pairwise deletion. Imputation methods substitute missing values and outliers based on the observed data. Despite their widespread adoption, these methods are not without drawbacks, notably the risk of compromised generalizability and the potential for increased bias, due to their respective modeling assumptions. Although these methods exist to ameliorate, they neglect the informative potential of missing data, particularly in critical settings like the Intensive Care Unit (ICU).

Similarly, the concept of sampling frequency is often overlooked in statistical analyses or predictive models, despite its critical importance, especially in the ICU. Sampling frequency refers to how often data points, such as vital signs or laboratory values, are collected. As noted by Zhang et al. [2021], the frequency of vitals collected can vary, and incorporating this variability can improve the performance of outcome prediction models. However, this sampling frequency is rarely explicitly considered. For example, studies such as those by Zhang et al. [2020] and another by Khope and Elias [2023] developed predictive models using the MIMIC-III dataset without incorporating sampling frequencies, instead focusing on static and aggregated temporal data. Ignoring informative sampling frequencies can significantly affect the quality and reliability of predictive models and statistical inferences.

Certain demographic groups may receive more frequent and thorough documentation due to various factors such as socioeconomic status, healthcare access, or underlying health disparities. These disparities in data capture can inadvertently introduce or amplify implicit biases in healthcare research, potentially leading to inequitable healthcare outcomes. Therefore, it becomes important to understand how demographic variables such as age, gender, and race/ethnicity might independently influence the presence of missing data and the sampling frequency of variables collected in EHRs.

This study introduces an analytical framework designed to measure the occurrences of missing data and the frequency of sampling in Electronic Health Records (EHRs). We apply this framework to the MIMIC-III dataset, the most frequently used EHR dataset in machine learning research and one of the very few publicly available datasets where protected attributes, such as demographic variables, are collected and documented (Fong et al. [2023]). Specifically, we propose to investigate the potential association between missing data and variability sampling frequencies and demographic variables, including age, gender, and race/ethnicity. We focus on sampling frequency and missing rate for key clinical and biological parameters in critical care patients. As a secondary goal, we demonstrate that variations in sampling frequency and missingness rates are predictive of in-hospital mortality.

## 2 Methods

### 2.1 Data Source

The data used in this study were sourced from the Medical Information Mart for Intensive Care III (MIMIC-III) Johnson et al. [2016]. This electronic repository contains patient-specific healthcare information and encompasses a variety of variables, including patient demographics, hospital mortality rates, diagnostic data, laboratory test results, prescription records, and medical procedures. The MIMIC-III database represents a cohort of more than 40,000 deidentified patients who were admitted to intensive care units (ICU) at Beth Israel Deaconess Medical Center in Boston, Massachusetts, from 2001 to 2012, and is publicly accessible through PhysioNet, subject to a data use agreement (Goldberger et al. [2000]).

For the purpose of this study, we included the first admission records of patients meeting the following eligibility criteria: (1) adult age, defined as 18 years or older; (2) absence of ICU admission related to pregnancy, childbirth, or the postpartum period; (3) no live discharge within the initial 24-hour period following admission; and (4) the presence of at least one arterial line, commonly used in ICUs for continuous blood pressure monitoring and arterial blood sampling, during the ICU stay (Shiver et al. [2006]).

### 2.2 Variables and Data Structure

In the ICU, the monitoring of acutely ill patients is conducted through a multiplicity of methods, including both laboratory tests (LTs) and vital sign measurements (VSs). Within the first 24 hours after admission to the ICU, it is standard practice to perform a range of laboratory tests (Ezzie et al. [2007]). In addition, the healthcare team continuously monitors and records vital signs. The frequency of these assessments and their entry into the EHR can vary according to the specific clinical requirements of each patient.

We extracted data pertaining to 11 distinct vital signs and 35 different laboratory tests, which were selected based on a list comprising the top 80% of commonly performed tests Frassica [2005] (see Appendix A, Table **??**). This data set also includes baseline characteristics for each patient and severity scores for each ICU stay(Johnson et al. [2018]).

To assess the influence of demographic variables on measurements and patterns of data missing, we extracted patient data from the initial 24-hour period of admissions to the ICU. Furthermore, to evaluate their dynamic changes and correlation with patient outcomes, we extended our data extraction to include information from the first five days of ICU stays, segmented into consecutive 12-hour intervals. Detailed information regarding the structure of the data is provided in Appendix B.

### 2.3 Statistical Analysis

#### 2.3.1 Measurement frequencies and missingness patterns

Two distinct types of measurement rate variables were used. The first type quantifies the total number of measurements per variable during the initial 24 hours or during subsequent 12-hour blocks. The second type, termed the missingness rate, quantifies the frequency of missing observations for each variable, calculated as the number of hours without any observation. For the first 24-hour block, this rate can range from 0 to 24, and for the 12-hour blocks, from 0 to 12. A detailed description of how these measurement rate variables were computed is provided in Appendix C.

Since some vital signs are often monitored together, we grouped them and consolidated their missingness and measurement rates into average rates. Similarly, for laboratory tests that are often ordered together, we grouped them and consolidated their measurement rates into single variables that reflect group averages, as detailed in Appendix C, Table **??**.

#### 2.3.2 Association of demographic variables and measurement rates

To estimate the association between demographic variables and measurement rate variables, we used the double robust Targeted Maximum Likelihood Estimator (TMLE) (Van der Laan and Rose [2018]). The TMLE framework is a versatile method for deriving efficient estimators of estimands in nonparametric models, suitable for applications such as measuring intervention impacts or deriving nonparametric variable importance measures using machine learning. Additionally, TMLE allows for robust (model-free) statistical inference.

We defined the observed data as *O_i_* = (*W_i_, A_i_, Y_i_*), *i* = 1*, …, n*, where *i* indexes the individual, *W_i_* are adjustment variables including health status and demographic factors, *A_i_* is the demographic variable of interest, and *Y_i_* is the measurement patterns outcome. Our parameter of interest can be thought of as the marginally adjusted mean: *θ*(*a*) = E E[*Y |A* = *a, W*], for each level a of the current demographic variable of interest. For example, comparisons such as *θ*(*Hispanic*) vs. *θ*(*White*) adjust for variables including health status variables such as the Sequential Organ Failure Assessment (SOFA) score and the other demographic variables. If *A* is defined as age, then *W* will contain both health status and additional demographic variables such as race.

TMLE provides an unbiased estimator of the target estimand if one consistently estimates the outcome regression *Q*(*A, W*) = E[*Y |A, W*] or the propensity score *g*(*a|W*) = *P* (*A* = *a|W*). Due to the lack of knowledge of the underlying models in observational studies, data-adaptive estimation methods are essential. We used a Super Learning (SL) ensemble machine learning algorithm with a library of both simple and highly adaptive (machine) learning to estimate both *Q* and *g*. The goal of using the combination of the parameter (estimand) and estimating method is to provide non-parametric, apples-to-apples comparisons of the mean measurement rates per demographic group, which aggressively adjust for clinical and other variables, and also provides robust statistical inference in the context of using highly adaptive machine learning algorithms. We conducted the analyses using the sl3 (SuperLearner) and tmle3 (TMLE) packages in the R language. [Coyle [2021]].

In addition to the primary analysis, we performed sensitivity analyzes using more traditional regression approaches (generalized linear models or GLMs). These analyses served to examine the coefficients of the demographic variables under different model specifications for predicted outcomes including missingness patterns and sampling frequencies. Initially, we excluded severity scores and other demographic variables to understand the unadjusted relationships, and subsequently included only severity scores as confounding factors. This multifaceted approach provided a comprehensive understanding of how demographic variables influence both measurement patterns and data completeness in critical care environments.

#### 2.3.3 Predictive power analysis

To explore the relationship between measurement pattern variables and patient outcome, we evaluated the predictive capacity of measurement-related variables alongside recorded clinical values to forecast ICU mortality in the next 12 hours. This evaluation employed the Super Learner (SL) algorithm, renowned for its ensemble method that enhances prediction accuracy through V-fold cross-validation and integrates a diverse range of predictive models, from simple linear regressions to complex machine learning techniques (Van der Laan et al. [2007]). We implemented discrete SL using 10-fold cross-validation stratified by patient ID, which was crucial for managing the repeated measures characteristic of our dataset and ensuring distinct separation between training and validation sets. The candidate models included Generalized Linear Model (GLM), Bayesian GLM, Generalized Additive Model (GAM), Ridge Regression, Elastic-net Penalized Regression (ElasticNet), Lasso Regression, Random Forests, Gradient Boosting Machine, and Bayesian Additive Regression Trees.

We trained a predictive model of future (in the next 12 hours) ICU mortality using ‘values variables’, which comprised baseline characteristics (*W*), vital sign measurements (*V S*), and laboratory test values (*LT*). Then we trained another predictive model of future (in the next 12 hours) ICU mortality using ‘counting variables’, including counts of baseline information (*n*_*W*), vital sign sampling frequencies (*n*_*V S*), vital sign missingness rates (*h*_*m*_*V S*), and laboratory test frequencies (*n*_*LT*). Then we trained another one using a combined set of both ‘values’ and ‘counting’ variables. Then we evaluate and compare the predictive performances from the three models trained with different set of variables to see the variable importance of the set.

We developed three predictive models to forecast ICU mortality within the next 12 hours. The first model used ‘values variables’ as predictors, which included baseline characteristics (*W*), vital sign measurements (*V S*), and laboratory test values (*LT*). The second model employed ‘counting variables’ as predictors, consisting of counts of baseline information (*n*_*W*), vital sign sampling frequencies (*n*_*V S*), vital sign missingness rates (*h*_*m*_*V S*), and laboratory test frequencies (*n*_*LT*). The third model used the combination of both ‘values’ and ‘counting’ variables. We evaluated and compared the predictive performances of these three models using cross-validated measures of fit to determine the importance of each set of variables.

All analyses were performed using the R software, version 4.3.1 (2023-06-16).

## Results

### Patient Demographics

In the MIMIC-III database, comprising 46,520 patients, 23,426 met our inclusion criteria for the study. Of these, a low ICU mortality rate was observed, with only 464 patients (2.02%) experiencing mortality within the first 5 days of their ICU stay during their initial admissions. The demographic composition of the cohort is summarized in Table 1. 58.4% of the patients were male, and 50.7% identified as Christian. The majority (51.2%) were English speakers, and 70.9% were white.

**Table 1:** Patient demographics summary table.

|  | No ICU Mortality (N= 22962) | ICU Mortality (N=464) | Overall (N= 23426) |
| --- | --- | --- | --- |
| **Age** |  |  |  |
| Mean (SD) | 64.8 (17.2) | 67.0 (18.2) | 64.8 (17.2) |
| Median (IQR) | 66.0 (23.0) | 69.0 (25.0) | 66.0 (23.0) |
| **Gender** |  |  |  |
| Female | 9531 (41.5%) | 210 (45.3%) | 9741 (41.6%) |
| Male | 13431 (58.5%) | 254 (54.7%) | 13685 (58.4%) |
| **Religion** |  |  |  |
| Christian | 11671 (50.8%) | 198 (42.7%) | 11869 (50.7%) |
| Not Christian | 3509 (15.3%) | 56 (12.1%) | 3565 (15.2%) |
| Missing | 7782 (33.9%) | 210 (45.3%) | 7992 (34.1%) |
| **Partner** |  |  |  |
| Partner | 11576 (50.4%) | 208 (44.8%) | 11784 (50.3%) |
| No Partner | 9869 (43.0%) | 180 (38.8%) | 10049 (42.9%) |
| Missing | 1517 (6.6%) | 76 (16.4%) | 1593 (6.8%) |
| **Language** |  |  |  |
| English | 11798 (51.4%) | 206 (44.4%) | 12004 (51.2%) |
| Not English | 1879 (8.2%) | 46 (9.9%) | 1925 (8.2%) |
| Missing | 9285 (40.4%) | 212 (45.7%) | 9497 (40.5%) |
| **Ethnicity** |  |  |  |
| White | 16300 (71.0%) | 306 (65.9%) | 16606 (70.9%) |
| Black | 1487 (6.5%) | 25 (5.4%) | 1512 (6.5%) |
| Hispanic | 674 (2.9%) | 17 (3.7%) | 691 (2.9%) |
| Asian | 497 (2.2%) | 14 (3.0%) | 511 (2.2%) |
| Other | 653 (2.8%) | 13 (2.8%) | 666 (2.8%) |
| Missing | 3351 (14.6%) | 89 (19.2%) | 3440 (14.7%) |
| **Insurance** |  |  |  |
| Yes | 22716 (98.9%) | 449 (96.8%) | 23165 (98.9%) |
| No | 246 (1.1%) | 15 (3.2%) | 261 (1.1%) |

### Differential Monitoring Patterns

Our analysis first focused on the initial 24 hours of ICU data, comprising 24,517 ICU stays. We observed an expected positive correlation between the severity of the patient’s condition, as indicated by the SOFA score, and the frequency of monitoring. The complete correlation plots of all demographic variables and measurement rates are available in Appendix D.

We observed significant differences in our analysis of monitoring patterns across different age groups, adjusting for health severity scores and other demographic variables. Elderly patients, particularly those in the 46-65 and above 65 age brackets, were more frequently monitored for vital signs than younger patients (Figure 1). For example, the adjusted average number of temperature measurements within the first 24 hours was 11.9 (95% CI: 11.7 to 12.1) for the 46-65 age group and 12.2 (95% CI: 12.0 to 12.4) for those over 65. In stark contrast, the 18-30 age group had a lower adjusted average count of 9.7 measurements (95% CI: 9.4 to 10.1), and the 31-45 age group had an average of 10.25 (95% CI: 9.9 to 10.6). Interestingly, the Glasgow Coma Scale (GCS) measurements did not follow this trend and were consistently collected across all age groups. In contrast with vital signs, a monotonic pattern was noted in laboratory tests, where the frequency decreased linearly with increasing age, consistent across all lab tests even after adjusting for SOFA scores and other covariates. Complete results, consistent across all sensitivity analyses, are provided in Appendix E.

**Figure 1:**
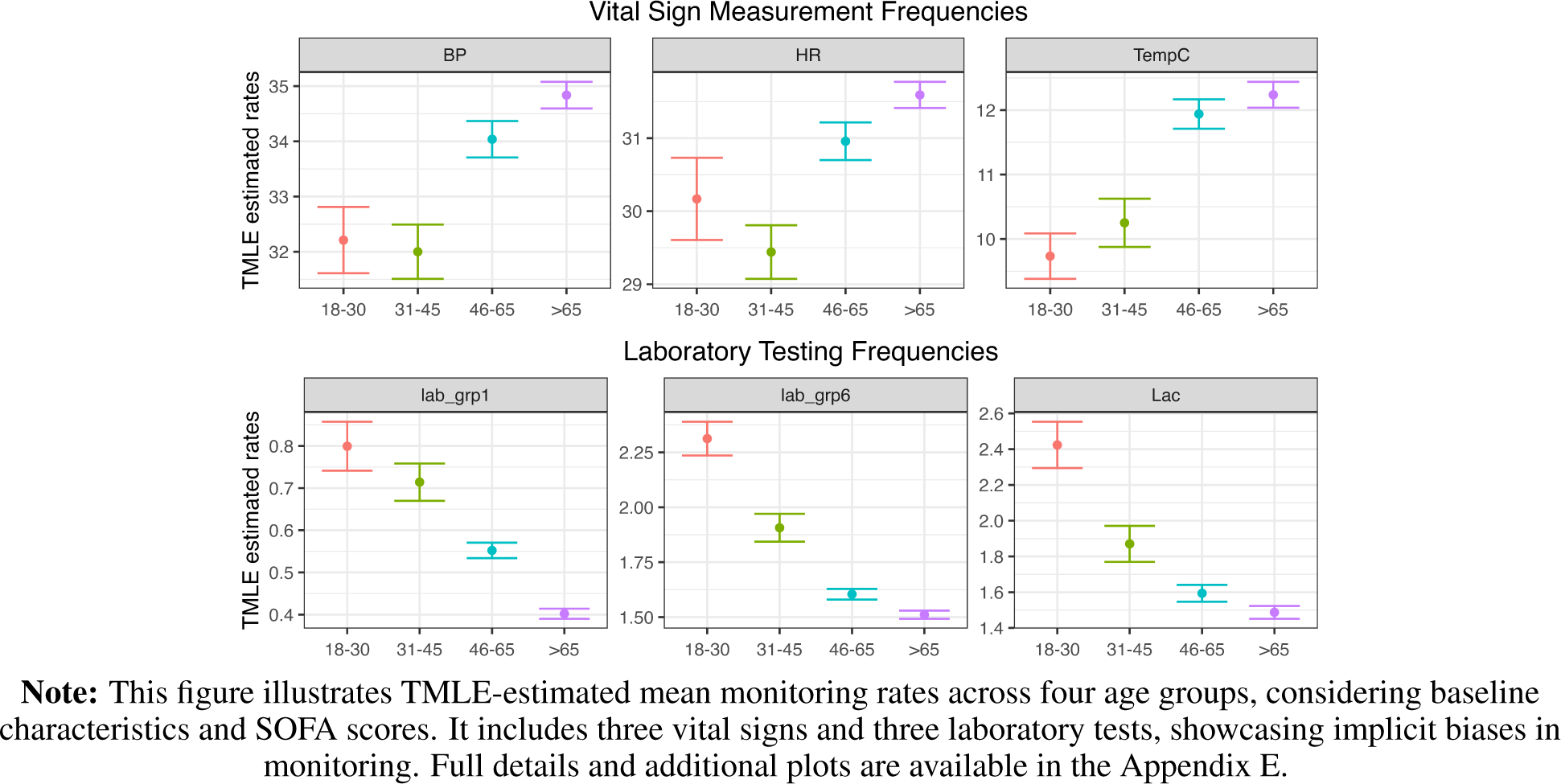
TMLE Estimated Monitoring Patterns by Age Groups.

**Figure 2:**
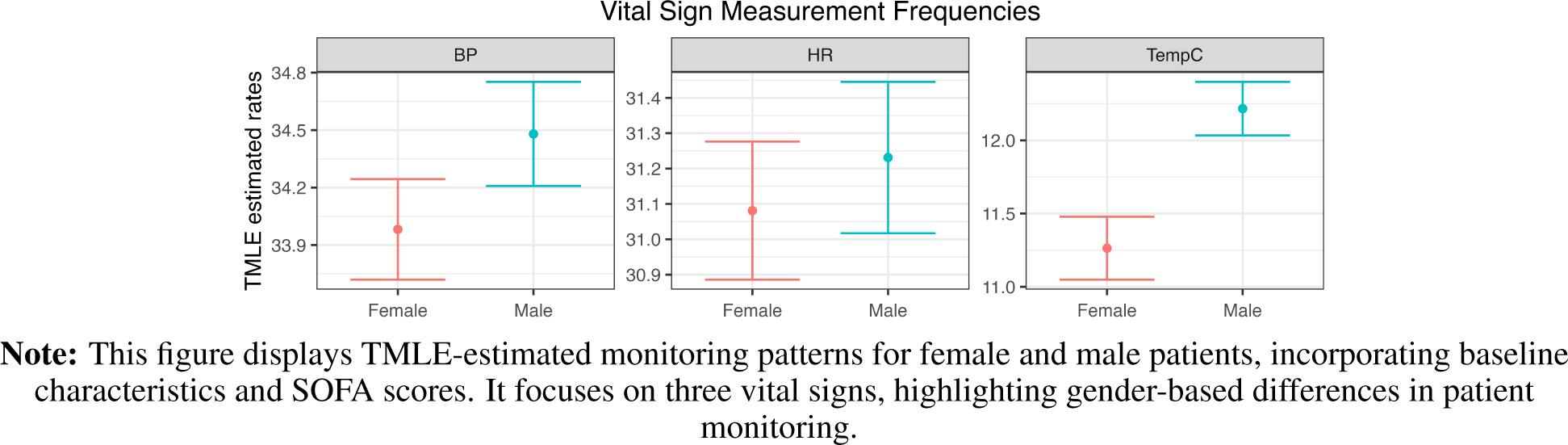
TMLE Estimated Monitoring Patterns by Gender.

Regarding gender, no significant differences were found across all vital signs (Figure 1), though males received marginally one more temperature checks in the first 24 hours: 12.22 (95% CI: 12.03 to 12.40) vs. 11.26 (95% CI: 11.05 to 11.50) temperature measurements.

Ethnicity was associated with monitoring patterns. We noted minor yet statistically significant disparities in the frequency of vital sign measurements between Black, Hispanic, and White groups, with the Black and Hispanic groups having fewer vital signs recorded. The adjusted average number of Blood Pressure measuring/recording in the first 24 hours, along with their 95% confidence intervals, were as follows: 34.22 (34.00, 34.44) for White, 34.45 (33.68, 35.22) for Asian, 32.79 (32.11, 33.46) for Black, 32.39 (31.82, 32.98) for Hispanic, and 33.74 (32.88, 34.59) for other race/ethnicities. The p-values for comparisons between White and Black, and White and Hispanic were 7.77e-05 and 8.09e-09, respectively.

For heart rate measurements, the adjusted averages were 31.09 (30.90, 31.24) for White, 31.15 (30.56, 31.75) for Asian, 30.11 (29.61, 30.60) for Black, 29.63 (29.19, 30.06) for Hispanic, and 30.79 (30.80, 30.15) for other ethnicities. Additional data on respiratory rate, oxygen saturation, and temperature measurements are detailed in Figure 3.

**Figure 3:**
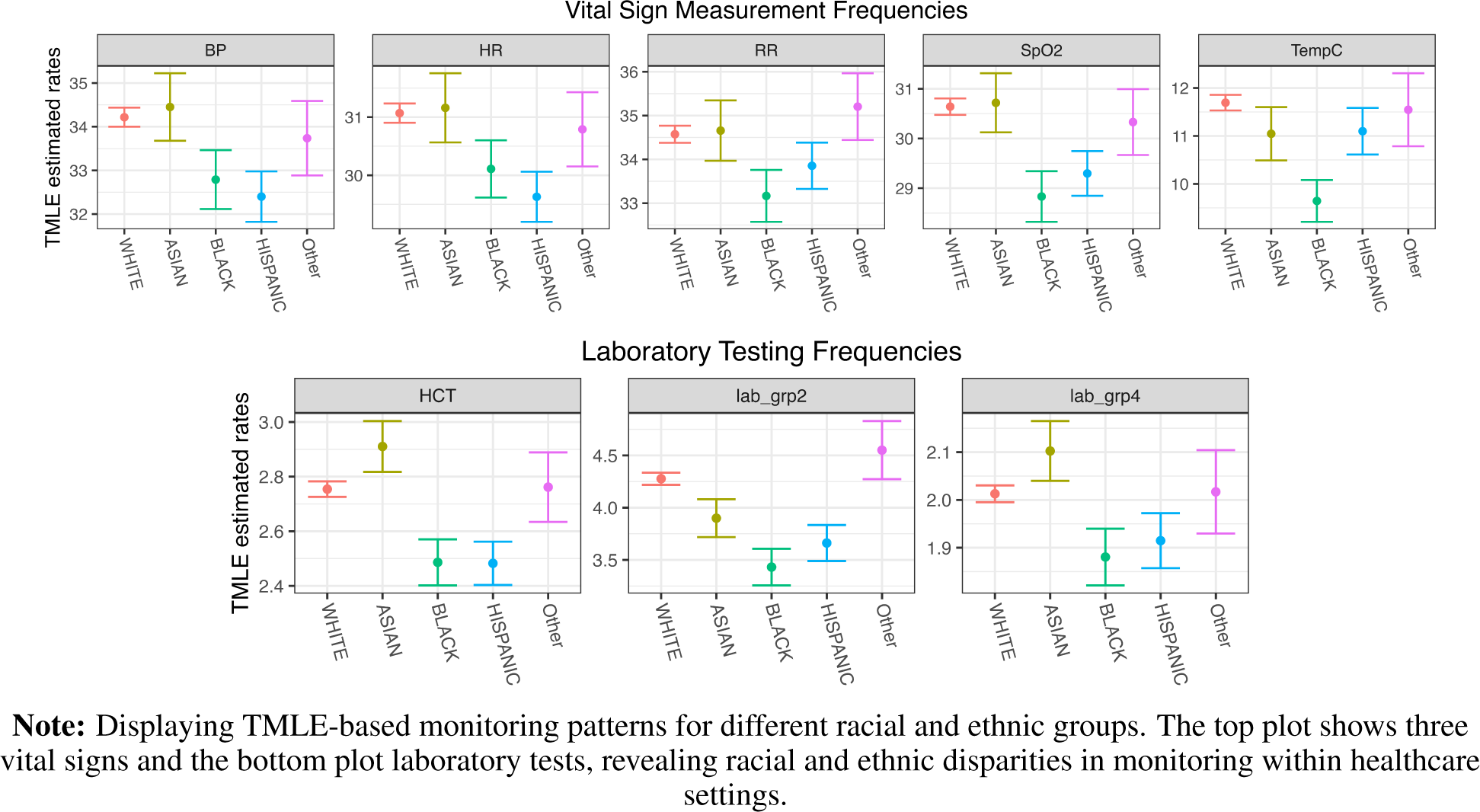
TMLE-Estimated Monitoring Patterns by Race/Ethnicity.

In contrast, laboratory tests did not show a consistent pattern of disparities, although some differences were observed in specific tests such as hematocrit levels. The adjusted average numbers with 95% confidence intervals for hematocrit were 2.49 (2.40, 2.57) for Black, 2.48 (2.40, 2.56) for Hispanic, and 2.75 (2.73, 2.78) for White.

Other demographic variables, including insurance status, language, marital status, and religion, showed no significant patterns in monitoring frequency. These findings are included in Appendix E.

### Measurement patterns predictive of mortality

Tables 4 and 2 report the fit metrics for different combinations of predictors, which include the measured and corresponding measurement rate and missingness variables. The model with the best fit was trained on the combination of the generated counting variables and the original variables. The performance of this fit was closely followed by the model trained solely on the original variables. The model exclusively trained on generated measurement rate/missingness variables, although slightly worse performing, still achieved impressive predictive accuracy. These results suggest that measurement rate variables alone can accurately predict mortality.

**Table 2:** Performance Metrics of Random Forest Models Across Different Variable Combinations.

|  | NLL | AUC | AUCPR |
| --- | --- | --- | --- |
| W, VS, LT | 0.17 (0.16, 0.19) | 0.85 (0.83, 0.87) | 0.18 (0.16, 0.20) |
| n_W, n_VS, h_m_VS, n_LT | 0.21 (0.18, 0.23) | 0.76 (0.74, 0.77) | 0.10 (0.09, 0.11) |
| W, VS, LT, n_W, n_VS, h_m_VS, n_LT | 0.17 (0.15, 0.19) | 0.86 (0.84, 0.87) | 0.19 (0.17, 0.22) |

**Note:** This table summarizes the average Negative Log-Likelihood (NLL), Area Under the ROC Curve (AUC), and Area Under the Precision-Recall Curve (AUCPR) for Random Forest models trained with different variable sets, based on 10-fold cross-validation.

The performance of included candidate learners in the SuperLearner library are presented in Table **??** in Appendix F. Figure 4 and Table 2 detail the performance metrics with 95% confidence intervals of Random Forest models, which demonstrated the best performance among the candidate learners through the ID-stratified 10 fold cross-validation process, where each RF models was trained using one of the three distinct sets of variables.

**Figure 4:**
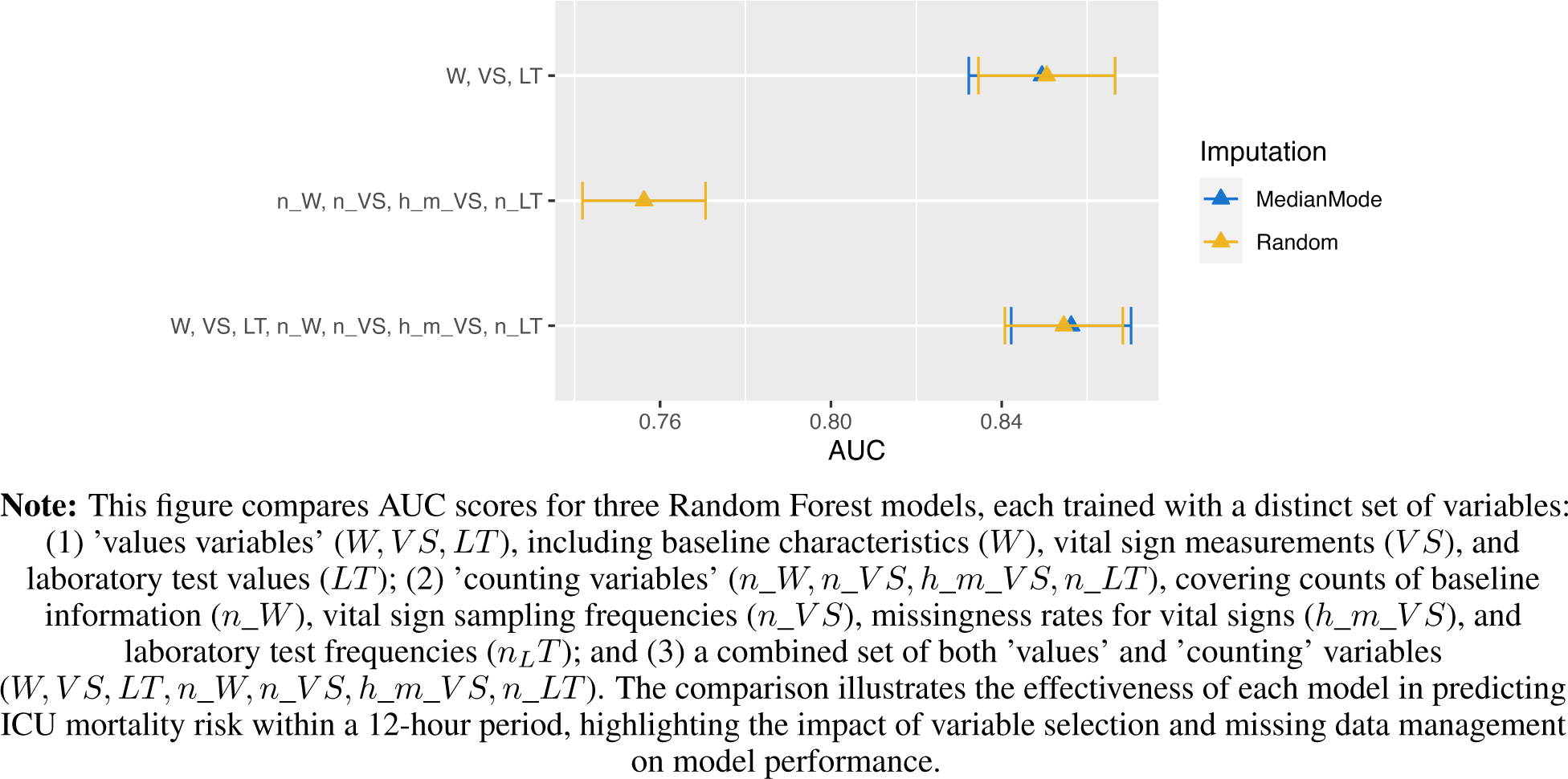
AUC Metrics in Random Forest Models Trained with Different Variable Combinations.

## 3 Discussion

### 3.1 Summary of Findings

We proposed a novel perspective on missing data and measurement frequencies within Electronic Health Records (EHR). Our analysis revealed statistically significant disparities in the sampling frequencies and missing data patterns in vital sign measurements and laboratory tests across different demographic groups in the first 24 hours. The older groups received more frequent monitoring with fewer data gaps compared to the younger groups, except in the Glasgow Coma Scale (GCS) measurements. Males received slightly more temperature checks in the first 24 hours. Notable disparities were found in the frequency of vital sign measurements between Black, Hispanic, and White groups, with the former groups contributing significantly fewer measurements. The frequency of laboratory test sampling decreased linearly with age, and while no consistent patterns of disparities were observed between other demographic groups, some differences were noted in specific tests such as hematocrit. We also examined the effects of insurance, language, marital status, religion, and care unit on data collection practices, but no strong patterns emerged.

Our analysis also demonstrated the strong predictive power of missing data patterns and measurement frequencies to patient outcomes in ICU settings. These variations in data collection practices can significantly affect the predictive accuracy of models that assess in-hospital mortality risks.

### 3.2 Discussion and Comparison with Previously Published Literature

We contribute to the growing body of literature highlighting biases in healthcare data analytics. Landmark studies, such as Obermeyer et al. [2019] and Suresh and Guttag [2019], showed significant biases in medical algorithms and healthcare data, emphasizing the need for equitable data representation. Our findings align with previous research indicating the presence of systemic biases in EHR data collection (Verheij et al. [2018] and Pivovarov et al. [2014]). Our focus on patterns of missing data and measurement frequencies, their associations with demographics, and their predictive power provide more insight into the consequence of this systematic measurement variability.

Previous studies (Rusanov et al. [2014] and Goldstein et al. [2017]) have explored the implications of missing data and sampling biases in predictive modeling, highlighting the potential for these factors to skew results and perpetuate inequalities. Our research builds on these findings by specifically analyzing the demographic disparities in missing data patterns and measurement frequencies. We found statistically significant disparities in these patterns between demographic groups and demonstrated the potential to improve the accuracy of the predictive model by including these disparities in the training data.

The discrepancy in data collection among different demographic groups, as evidenced in our study, underscores a larger issue of inequality in the representation of healthcare data. To address the disparities in the collection of EHR data, we recommend incorporating not just advanced imputation techniques, but also a more in-depth analysis of sampling patterns into statistical models. Implement advanced imputation techniques such as multiple imputation by chain equations (MICE) and deep learning-based methods that can be tailored to account for the demographic characteristics of the data to ensure fairness (Samad et al. [2022]; Sun et al. [2023]). Second, and more importantly, including sampling patterns in statistical models and machine learning can mediate the impact brought by different sampling intentions. This can be achieved by weighting the samplings and observations according to their measurement frequencies, modeling the data generation process as a multilevel model where the sampling pattern is treated as a latent variable influencing the observed data, or as a Bayesian model estimating the impact of different sampling frequencies on model outcomes, providing a probabilistic framework to handle uncertainty (Little and Rubin [2019]; Gelman and Hill [2006]; Gelman et al. [1995]; Rusanov et al. [2014]).

Finally, promoting transparency in data collection processes and conducting regular audits of EHR systems to detect bias can help create a more equitable healthcare data landscape. Our comparison with previously published studies underscores the continued need for methodological advancements to tackle these complex challenges and ensure that healthcare analytics are fair and accurate (Obermeyer et al. [2019]; Suresh and Guttag [2019]).

### 3.3 Strengths and Limitations of Our Study

One of the primary strengths of our study is its innovative approach to analyzing EHR data, particularly in identifying and interpreting biases in missing data and measurement frequencies. Our methodology provides a framework for future research in this area. However, limitations include the potential generalizability of our findings, as our study was confined to the MIMI-III database. Additionally, the retrospective nature of the study may limit the ability to infer causality between data collection practices and patient outcomes.

### 3.4 Conclusion

In conclusion, our study highlights the critical need for a sophisticated and equitable approach to handling missing data and measurement frequencies when using EHR data, especially in the EHR settings. By analyzing patterns of missing data and measurement frequencies, we can gain insights into patient monitoring practices and healthcare equality. Incorporating these insights can enhance predictive models in terms of both accuracy and inclusiveness. This advancement not only propels the field of healthcare data analytics forward but also contributes to the development of a more equitable healthcare system. Future research should assess the bias introduced by measurement patterns in the predictive algorithms and develop a general method to incorporate these patterns in the training data, thereby improving the reliability and inclusiveness of healthcare data analytics and algorithms.

## Data Availability

All data produced in the present study are available upon reasonable request to the authors

---

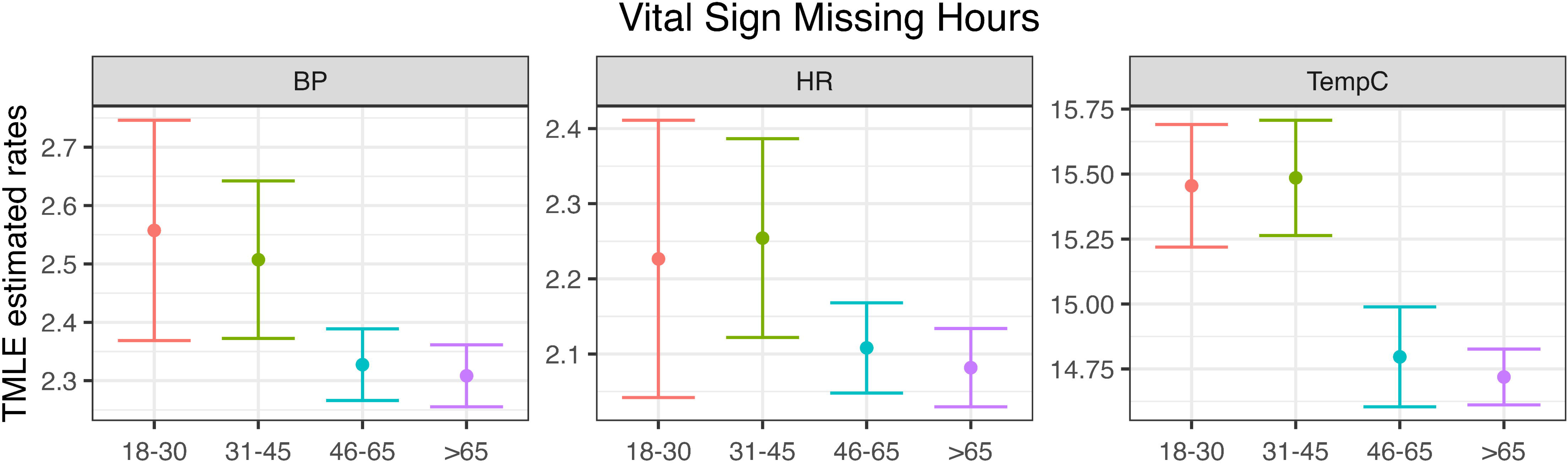

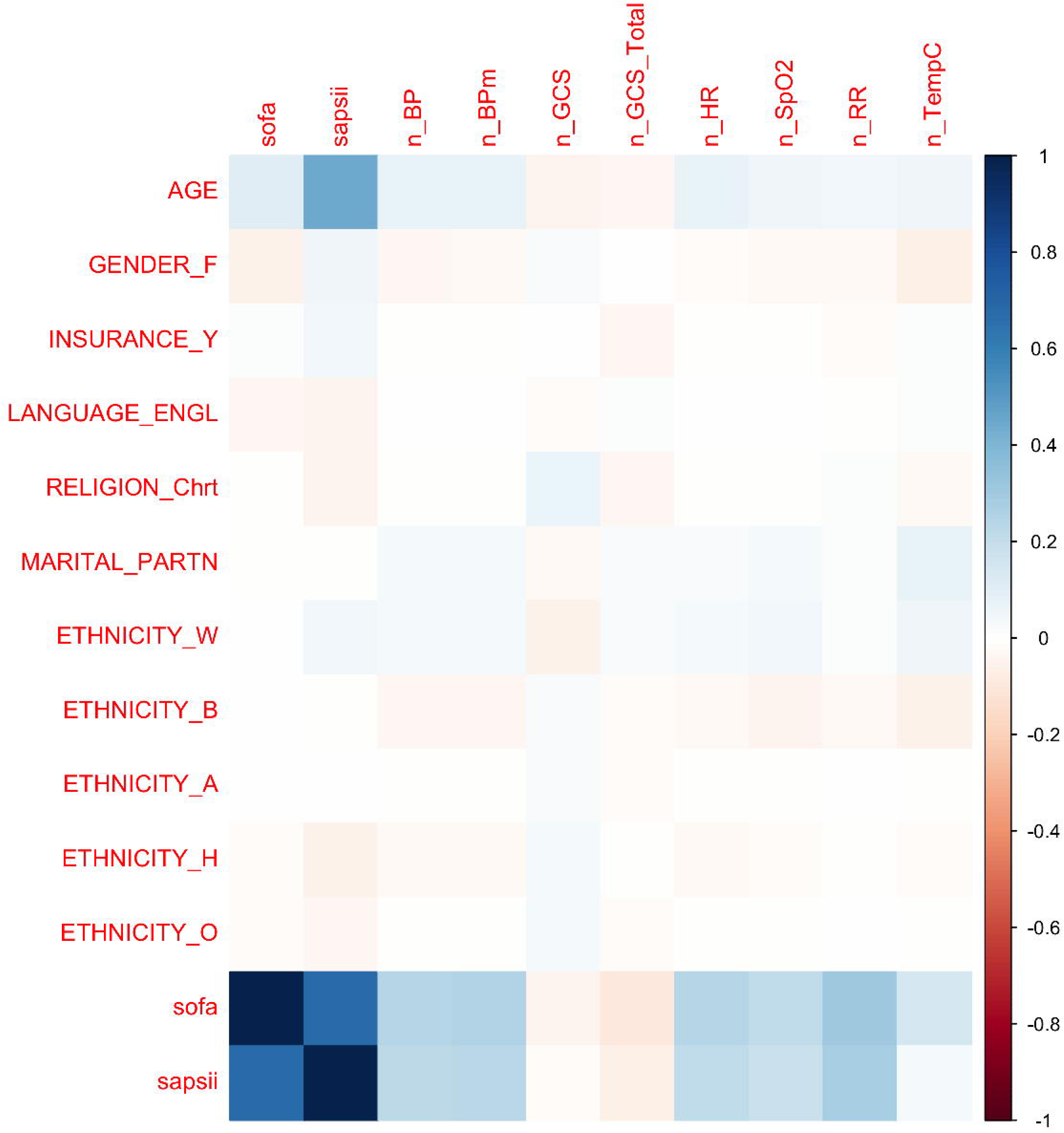

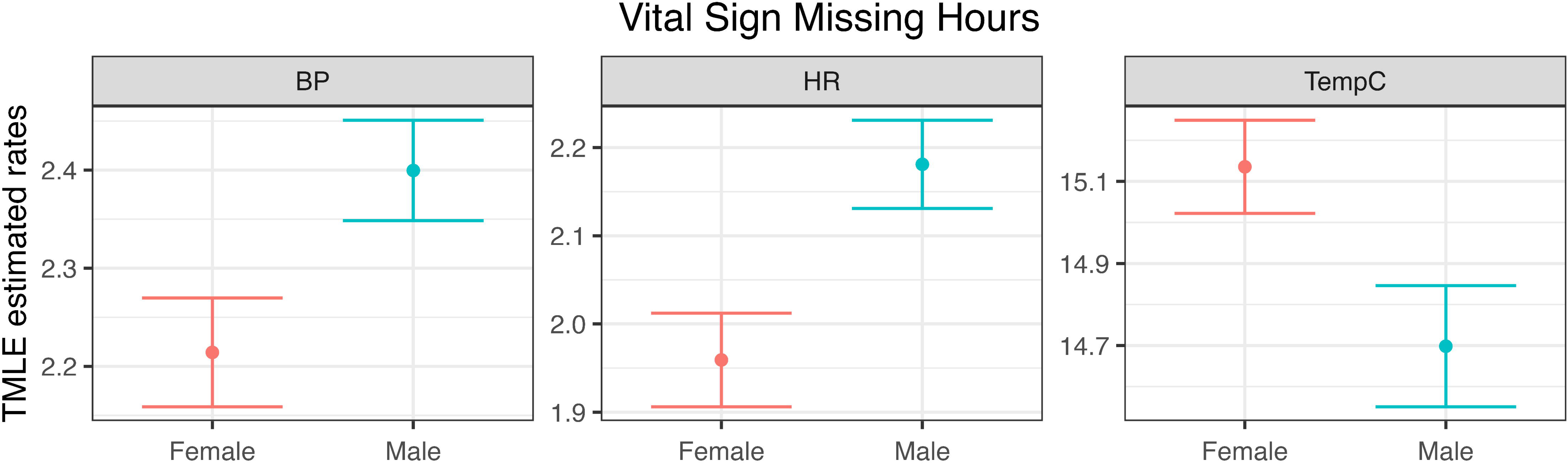

